# Metagenomic sequencing to detect respiratory viruses in persons under investigation for COVID-19

**DOI:** 10.1101/2020.09.09.20178764

**Authors:** Ahmed Babiker, Heath L. Bradley, Victoria D. Stittleburg, Autum Key, Colleen S. Kraft, Jesse J. Waggoner, Anne Piantadosi

## Abstract

We used metagenomic next-generation sequencing (mNGS) to assess the frequencies of alternative viral infections in SARS-CoV-2 RT-PCR negative persons under investigations (PUIs) (n=30) and viral co-infections in SARS-CoV-2 RT-PCR positive PUIs (n=45). mNGS identified both co-infections and alternative viral infections that were not detected by routine clinical workup.

## Introduction

The coronavirus disease 2019 (COVID-19) pandemic caused by severe acute respiratory syndrome coronavirus 2 (SARS-CoV-2) has precipitated a massive global health and economic crisis, claiming over a quarter million lives since its emergence in Wuhan, China in December, 2019 (1). Broad testing for respiratory viruses among persons under investigation for SARS-CoV-2 is performed inconsistently, limiting our understanding of alternative viral infections and co-infections in these patients. We evaluated the performance of metagenomic next-generation sequencing (mNGS) to detect SARS-CoV-2, co-infections, and alternative viral infections in persons under investigation (PUIs) for SARS-CoV-2 during the first two months of the pandemic in the state of Georgia.

## Materials and Methods

A convenience sample set was selected from 75 PUIs who were tested for SARS-CoV-2 in the Emory Healthcare system between February 26^th^ and April 23^rd^ 2020 (spanning the first detection of SARS-CoV-2 infection in Georgia on March 2^nd^ and in the Emory Healthcare system on March 3^rd^). Clinical data including laboratory results were extracted by chart review. Flu/RSV PCR (Cepheid, Sunnyvale, CA), and multiplex respiratory panels including the eSensor® Respiratory Viral Panel (GenMark Diagnostics, Inc., Carlsbad, CA), BioFire® FilmArray® Biofire Respiratory pathogen and BioFire® FilmArray® Pneumonia panel (BioFire Diagnostics, LLC, Salt Lake City, UT) had been performed at the discretion of treating physicians. Excess nasopharyngeal swab samples were retrieved within 72 hours of collection and underwent RNA extraction and SARS-CoV-2 testing by triplex RT-PCR (2). Samples underwent unbiased mNGS as previously described (3, 4). Briefly, this included DNAse treatment, random primer cDNA synthesis, Nextera XT tagmentation, and Illumina sequencing to a median depth of 35 million reads. Reads underwent taxonomic classification by KrakenUniq, as implemented in viral-ngs (5). Reads that were taxonomically assigned to a human respiratory virus were extracted, aligned to a reference sequence in Geneious Prime V2020.1.1, and manually inspected. Detection of a virus was reported as confirmed (**Supplementary Tables 1 and 2**) if nonoverlapping reads from ≥3 distinct genomic regions were identified (6). Reference-based viral genome assembly was performed using viral-ngs. The following references were used for alignment and assembly: NC_001803.1 (Respiratory Syncytial Virus (RSV)), NC_0391991.1 (Human Metapneumovirus), NC_006577.2 (Human Coronavirus (HCoV) HKU1), NC_005831 (HCoV NL63), NC_006213.1 (HCoV OC43), and NC_026431.1-NC_026438.1 (Influenza H1N1). In some cases, reads were assigned to a virus by KrakenUniq but failed to align to the reference; these reads underwent classification by BLASTn. All reads (cleaned of human reads) will be submitted to the Sequence Read Archive and complete genomes will be submitted to GenBank.

## Results

75 PUIs were included, 30 of whom were negative and 45 of whom were positive for SARS-CoV-2 by RT-PCR. Among the 30 SARS-CoV-2 negative PUIs, 93% (28/30) underwent clinical testing for alternative infections, 73% (22/30) by Flu/RSV PCR testing and 80% (24/36) by multiplex respiratory viral panel testing. Eight of the 30 SARS-CoV-2 negative PUIs (27%) tested positive for another respiratory virus by routine clinical testing. In all cases, mNGS identified the same virus (**Table 1**). Importantly, mNGS also identified four viruses that were not detected by routine clinical testing: two (Metapneumovirus and HCoV NL63) that were not tested for and two (RSV and influenza, in the same patient sample) that were missed despite testing with a multiplex respiratory viral panel (FilmArray Biofire Respiratory pathogen). In addition to these confirmed findings, mNGS identified one read of HCoV NL63 in an otherwise negative sample, which was below the limit of detection for this study, as well as reads initially classified as alphacoronaviruses in 3 samples, which were ultimately determined to be non-viral contaminants by BLASTn. No SARS-CoV-2 was detected by mNGS among RT-PCR negative PUIs (**Supplementary Table 1**).

**Table 1:**
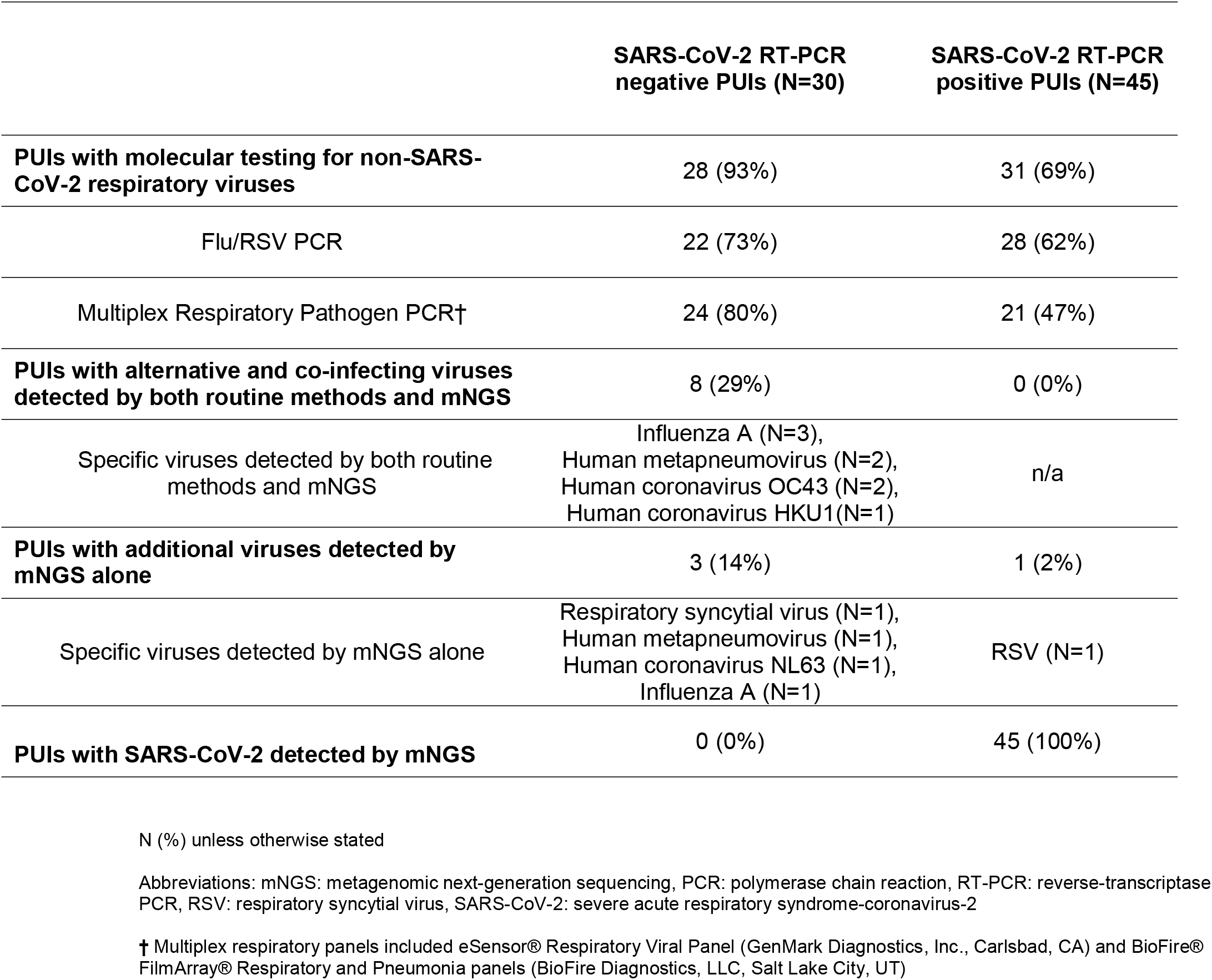
Molecular and Metagenomic Testing of Persons Under Investigation.

|  | SARS-CoV-2 RT-PCR<br>negative PUIs (N=30) | SARS-CoV-2 RT-PCR<br>positive PUIs (N=45) |
| --- | --- | --- |
| **PUIs with molecular testing for non-SARS-CoV-2 respiratory viruses** | 28 (93%) | 31 (69%) |
| Flu/RSV PCR | 22 (73%) | 28 (62%) |
| Multiplex Respiratory Pathogen PCR† | 24 (80%) | 21 (47%) |
| **PUIs with alternative and co-infecting viruses detected by both routine methods and mNGS** | 8 (29%) | 0 (0%) |
| Specific viruses detected by both routine methods and mNGS | Influenza A (N=3),<br>Human metapneumovirus (N=2),<br>Human coronavirus OC43 (N=2),<br>Human coronavirus HKU1(N=1) | n/a |
| **PUIs with additional viruses detected by mNGS alone** | 3 (14%) | 1 (2%) |
| Specific viruses detected by mNGS alone | Respiratory syncytial virus (N=1),<br>Human metapneumovirus (N=1),<br>Human coronavirus NL63 (N=1),<br>Influenza A (N=1) | RSV (N=1) |
| **PUIs with SARS-CoV-2 detected by mNGS** | 0 (0%) | 45 (100%) |

N (%) unless otherwise stated
Abbreviations: mNGS: metagenomic next-generation sequencing, PCR: polymerase chain reaction, RT-PCR: reverse-transcriptase PCR, RSV: respiratory syncytial virus, SARS-CoV-2: severe acute respiratory syndrome-coronavirus-2
† Multiplex respiratory panels included eSensor® Respiratory Viral Panel (GenMark Diagnostics, Inc., Carlsbad, CA) and BioFire® FilmArray® Respiratory and Pneumonia panels (BioFire Diagnostics, LLC, Salt Lake City, UT)

Among the 45 SARS-CoV-2 RT-PCR positive PUIs, 31 (69%) underwent clinical testing for co-infections, and none were detected (**Table 1**). mNGS identified one viral co-infection (RSV) that was not tested for; this was confirmed with a second independent sequencing library. SARS-CoV-2 was detected by mNGS in all 45 RT-PCR positive PUIs (**Supplementary Table 2**).

## Discussion

Overall, using unbiased mNGS, we were able to readily identify SARS-CoV-2, co-infections, and alternative infections in PUIs, including viruses not identified by routine clinical testing. Reassuringly, we did not detect SARS-CoV-2 by mNGS in samples that were negative by RT-PCR, indicating both that the RT-PCR assay was sensitive, and that there were no divergent viruses missed due to primer mismatch (7).

Interestingly, many PUIs had no infection identified on either routine testing or mNGS, which may reflect inadequate sampling, rapid clearance of a potential viral infection, or a non-viral process. Limitations of our study include the testing of banked samples which may degrade over time with freeze-thaw cycles, and the use of clinical tests (such SARS-CoV-2 RT-PCR) as an imperfect gold standard. Nevertheless, our results offer several valuable insights regarding molecular testing for respiratory viruses among SARS-CoV-2 PUIs.

First, we found a low rate of viral co-infection in patients with SARS-CoV-2, with only one co-infection (with RSV) among 45 patients (2%). Our results are concordant with a systematic review of 16 studies and 1,014 patients, which found a 3% rate of viral co-infection, with RSV and Influenza A being the most common (8). Thus, viral co-infections may be less common than initially reported (9); nevertheless, it is imperative to continue to monitor for them in the setting of the upcoming respiratory virus season and ongoing waves of COVID-19.

Second, our results illustrate key considerations about the potential use of mNGS as a diagnostic tool. Notably, mNGS detected all viruses that were identified by routine clinical workup, as well as five viruses that were not identified by clinical workup. Three of these five had not been tested for clinically, underscoring the importance of routine broad-spectrum molecular testing for respiratory viruses among PUIs. However, two of the viruses were tested for and not identified by multiplex PCR, suggesting that mNGS may offer improved sensitivity. Additional advantages of mNGS have been reviewed elsewhere (10) and include the opportunities to characterize viral genomics, the microbiome, and the host transcriptome.

However, current challenges with implementing mNGS into routine clinical microbiology workflows include its relatively high cost and low throughput, as well as the need for expertise in interpreting results. One barrier in particular is the lack of established thresholds for identifying pathogens. Herein, we required detection of nonoverlapping reads from ≥3 distinct genome regions based upon previously described criteria (6). Recently, Peddu *et al*. used a cutoff based on reads per million RPM (< 10) (11); in our study, the use of this cutoff would have excluded viruses that were found by both mNGS and routine clinical testing by PCR, which we regard as true positives. Thus, there is ongoing need to harmonize practices around clinical mNGS testing.

In conclusion, our results suggest that current broad-spectrum molecular testing algorithms identify most respiratory viral infections among SARS-CoV-2 PUIs, when available and implemented consistently. In addition, these results illustrate the potential of mNGS to streamline and expand clinical testing for respiratory viruses, which may augment strategies to surveil for unexpected viral co-infections or the emergence of divergent strains during periods of high transmission.

## Data Availability

All reads (cleaned of human reads) will be submitted to the Sequence Read Archive and complete genomes will be submitted to GenBank.

## Funding

This study was supported by the Pediatric Research Alliance Center for Childhood Infections and Vaccines and Children’s Healthcare of Atlanta. The Yerkes NHP Genomics Core is supported in part by NIH P51 OD011132 and sequencing data was acquired on an Illumina NovaSeq6000 funded by NIH S10 OD026799. Salary support was provided by the Doris Duke Charitable Foundation, Clinical Scientist Development Award 2019089 (JJW) and NIH K08 AI139348 (AP).

## Acknowledgments

We would like to acknowledge our laboratory colleagues at the Emory University Healthcare Microbiology and Molecular laboratories who have worked tirelessly to provide necessary care to our patients during this time. We would also like to thank the staff of the Emory Integrated Genomics Core, Yerkes NHP Genomics Core and Jessica Ingersoll of the Emory Clinical Virology Research Laboratory for sequencing support, and Daniel J. Park for valuable advice on the use of viral-ngs.

## Notes

### Competing Interest Statement

The authors have declared no competing interest.

### Funding Statement

This study was supported by the Pediatric Research Alliance Center for Childhood Infections and Vaccines and Childrens Healthcare of Atlanta. The Yerkes NHP Genomics Core is supported in part by NIH P51 OD011132 and sequencing data was acquired on an Illumina NovaSeq6000 funded by NIH S10 OD026799. Salary support was provided by the Doris Duke Charitable Foundation Clinical Scientist Development Award 2019089 (JJW) and NIH K08 AI139348 (AP).

### Author Declarations

Emory University IRB

